# SEXUAL BEHAVIOURS AND EXPERIENCE OF SEXUAL COERCION AMONG IN-SCHOOL FEMALE ADOLESCENT IN SOUTHWESTERN NIGERIA

**DOI:** 10.1101/19000851

**Authors:** Babatunde Owolodun Samuel, R.A Sanusi

## Abstract

Most people begin their sexual relationship during adolescence and some get involved in risky life threatening behaviors such as unwanted pregnancies, abortions and sexually transmitted infections. This study was therefore designed to understand the patterns of female adolescents sexual behaviours and sexual coercion experience.

A descriptive cross-sectional study was carried out among 1227 in-school adolescents in the three senatorial district of Osun State, Southwestern Nigeria. Multi-stage sampling technique was used to select the respondents, and data were collected with pre-tested, semi-structured questionnaires. The data collected were analysed using Statistical Package for Social Sciences (SPSS version 21).

Three hundred and thirty seven (27.5%) were sexually exposed with a mean age of sexual initiation of 14.88±2. 46 years. Of the 337 that were sexually exposed 153(56.5%) initiated sex early between the ages of 10-15 years, while (4.7%) had uses drugs or take alcohol before sexual intercourse. Findings revealed that of those that were sexually exposed, 122(11%) were forced to have sex, 101(9.1%) played sex willingly, while 29(2.6%) felt threatened and 3(0.3%) were convinced with money. The proportion of the respondent who reported rape and abduction was (2.6%).

Finding from this study is consistent with earlier studies conducted in many other Nigerian cities which showed that in-school adolescents to be sexually active. There is the need to step up campaigns to address this noticed lapse in behavior among the students in order to arrest the usual consequences of such risky sexual behavior.

## INTRODUCTION

One of the fundamental human rights, according to the United Nations Education Scientific and Cultural Organization-UNESCO is the right to protection from harmful influences, abuse and exploitation (United Nations Children Emergency Fund, 1989). United Nations Children Emergency Fund-UNICEF reiterated that its mission is to advocate for the protection of children’s rights, especially children under the age of 18 years, to help meet their basic needs and to expand their opportunities to reach their full potentials. These rights among others include the right to protect children from sexual harassment and abuse, and right to be educated (United Nations Education, Scientific and Cultural Organization, 1978).

The period of adolescence is one of the most intriguing and difficult transitions in the life span of a mankind (Adegoke, 2003). Adolescence is defined as a period characterized by an series of challenges and confusion both to the adolescents and the adults who are supposed to show understanding (Moronkola, 2003). According to (Krost, *et.al*, 2001), the period of adolescence is the most controversial of all the three developmental stages due to experimental risky behaviors associated with it.

Most people begin their sexual relationship during adolescence and some get involved in risky life threatening behaviors such as unwanted pregnancies, abortions and sexually transmitted infections (Action Health Incorporated, 2003). This can be attributed to ignorance in interpreting and managing self in response to the upsurge of hormones during this period. Contributory factors also include lack of information on sexual health and HIV, low levels of condom use, and high levels of sexually transmitted infections (UNGASS, 2010)

Adolescents in Nigeria were seen as a healthy segment of the population and received low priority for services until recently. In Nigeria adolescent have high burden of reproductive health problems and this assertion is supported by surveys conducted on sexual behaviours of adolescents which shows that Nigerian adolescent (15-19) almost half of the females (46.2%) and about a quarter of males (22.1%) have engaged in sexual intercourse (National Demographic Health Survey, 2013; FMOH National HIV/AIDS and Reproductive Health Survey 2007; FMOH Integrated Biological and Behavioural Surveillance Survey, 2010).

Bothersome still is data from the Federal Ministry of Education in 2009 which found that 21% of upper primary school children surveyed indicated they have been involved in sexual intercourse yet only 40.6% who had two or more sexual partners in the past 12 months reported using a condom during their last sexual intercourse (National Demographic health Survey, 2008). A study conducted in Nigeria by WHO and United Nations Children’s Fund (UNICEF) revealed that about 2,400,000 and 2,600,000 people at 15 years of age had HIV/AIDS in 2003 and 2005, respectively (UNFPA, 2014).

Also in Nigeria, where the policy on population specifies the reduction of teenage pregnancy as a target to be met by the year 2010, the gravity of the problem is highlighted by result of the 2008 Demography and Health survey (DHS) which revealed that 23% of women aged 15 – 19 years are already mothers or are pregnant with their first child (NPC, 2009). According to the Federal Republic of Nigeria (FRON, 2012), twice as many girls than boys engage in sexual activity before the age of 15 years. Adolescents are exposed to high risk of sexually transmitted infections (Ergene, *et.al*., 2005). The situation becomes worse in a country like Nigeria where the concept of sex cannot be openly discussed even while the young people are clearly sexually active.

Female adolescents have sexual and reproductive health needs that remain poorly understood or adequately attended to worldwide (WHO, 2004). It can be seen from the foregoing that neglecting this population has serious implications on the future of any nation. Sexual activities of female adolescents predispose them to adverse effects including unwanted pregnancies, unsafe abortions and sexually transmitted diseases including HIV/AIDS. In Nigeria, both the brothel and non-brothel female sex workers rank first on the most at risk to HIV infections group (Federal Republic of Nigeria, 2012). According to (Wellings *et.al*., 2006), the use of condoms has increased among adolescents but levels of use are still not sufficient to substantially reduce the spread of HIV (Central Intelligence Agency World Fact book, 2010).

Sexual coercion is an increasingly common experience among female youth throughout the developing world and Several studies have suggested that the experience of sexual coercion leads to a greater likelihood of risky sexual behavior, such as early sexual debut, many sexual partners, and inconsistent condom use (Garoma *et.al*., 2008; Hovsepian *et.al*., 2010).Sexual coercion refers to an array of encounters that compel an individual to have sex against his or her will. These encounters include violence, threats of violence, verbal threats and insistence, deception, or economic conditions that leave an individual with a lack of an ability to choose not to engage in sex (Heise *et al*.,1995).

Sexual coercion among young people encompasses a range of experiences, ranging from noncontact forms such as verbal sexual abuse, kissing, caressing, petting, genital touching, attempted rape, forced penetrative sex (vaginal, oral, or anal), and any other sexually laden behavior that makes the victim feel uncomfortable. It also includes sex obtained as a result of intimidation, pressure, blackmail, deception, forced alcohol and drug use, and threats of abandonment or of withholding economic support. Transactional sex through money, gifts, or other economic incentives (especially in the context of extreme poverty) often has a coercive aspect as well (Shireen *et.al*., 2003; Michael *et.al*., 2004).

The consequences of sexual coercion are alarming: they are short and long-term; and have physical, psychological and social effects. School performance can also be affected. It has negative association with social and health outcomes, and it therefore, is an important public health issue. Significant number of young people experience sexual coercion (nonconsensual sex), such coercion is a violation of a person’s rights and can have severe physical, mental and reproductive health consequences including the risk of unintended pregnancy and HIV and other sexually transmitted infections (Shireen *et.al*., 2003; Michael *et.al*., 2004; WHO, 2004).

The level of sexual coercion has now become a matter of concern not only to guidance counselors, social workers but also those who are interested in the sexual reproductive health of adolescents and children. The preponderance occurrence of sexual coercion is noticeable recently by in Nigeria. Igbokure in Sunday Punch April 28, 2007 reported that some female students in the primary and secondary schools protested to the then Minister of education Dr. (Mrs) Ezekwesili over sexual harassment which they have been experiencing from their male teachers. The students urged the minister to let the male teachers know that the students came to the school to learn and not to be sexually harassed and intimated. Girls and women at all stages of their lifes are often the victims of sexual coercion. However it seems much of sexual coercion in Nigeria occurs to children and adolescents especially the females.

In many parts of the world, sexual coercion has been linked to risky sexual behavior, implying that there may be common causal pathways in the psychological reaction to serious violations of an individual’s integrity: The society believes that a male could force sex on female if he has spent a lot of money on her. Even if the woman reports any violence, the society tends to blame the female. It seems the economic crisis and poverty in Nigeria is making the female increasingly vulnerable to sexual coercion and making them to unable to resist pressure from old men who provide money and gift in exchange for sex. In some cases parents even force their female children to marry people who offer them money even if the marriage is against the girl’s wish.

In Nigeria, as elsewhere in Sub-Saharan Africa, studies confirm that a large proportion of adolescents have unmet reproductive health needs. Evidence of unmet need is reflected in the fact that some adolescents and other young person’s lack adequate knowledge and understanding of the reproductive process. Although, these researches have contributed to our understanding of the reproductive health behavior of young people but Public concern over the reproductive health problems among Nigerian youth is still drawing attention of researchers, non-governmental organizations (NGOs) and policy makers to examine the driving force behind the upsurge in adolescent sexual activity. The concern of this current research is to understand the patterns of female adolescents sexual behaviours and experiences of sexual coercion.

## METHODS

### Study Area

The study was carried out in Osun State. The State of Osun has three senatorial districts namely Osun Central, Osun West, and Osun East, each comprising 10 local government areas (LGAs) (making 30 LGAs) and one area office, located in Osun East senatorial district. The 30 LGAs are grouped into 18 rural and 12 urban LGAs. In the rural areas, majority of the inhabitants are farmers while in the urban areas, they are mostly traders, artisans, cloth dyers, and civil servants, hence having differing socio-economic status. The following Local Government area selected for collection of data were: Osun Central: Orolu, Irepodun, Osun East: Ilesa east, Ilesa west, Osun West: Ede north, Ede south

### Study Design

This was a descriptive cross-sectional study, carried out among 1227 in-school female adolescents in Osun State.

### Target Population

All of the adolescents in the area were the target population. Eligible students were 15–19 years old and who were presently enrolled in school.

### Sample Size Determination

Sample size was determined using Fischer’s sample size formula as shown below:

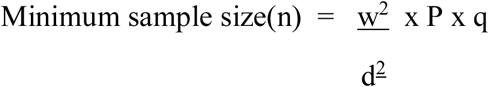

Where,

n= minimum sample size

w= standard normal value corresponding to 95% confidence level set at 1.96

d= level of error/tolerance 5%

p = P is the prevalence of sexual exposure 40% (from a previous study done among in school adolescents who had sexual initiation while still in secondary school in Nigeria (Otoide *et.al*., 2001)

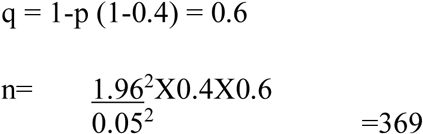

Adjusting the sample size for 10%attrition

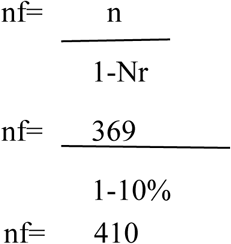

However, to increase representativeness in each of the senatorial districts and to improve the quality of the work, the sample size was tripled which added up to 1227

### Sampling Technique

Multi-stage sampling technique would be used.

Stage 1: Selection of six (6) Local Government Areas, two (2) from each of the three senatorial districts in Osun State through simple random sampling (with balloting with replacement). The following Local Government Areas were selected: Osun Central: Orolu, Irepodun, Osun East: Ilesa east, Ilesa west, Osun West: Ede north, Ede south

Stage 2: Selection of a ward from each of the local government areas using simple random sampling.

Stage 3: A list of Secondary schools in each local government was obtained from Osun State Ministry of Education. Schools was selected at random; one from each of the wards in each local government, selected.

Stage 4: From each the schools chosen, a proportionate sample was taken from each school by systematic random sampling using the formula below:

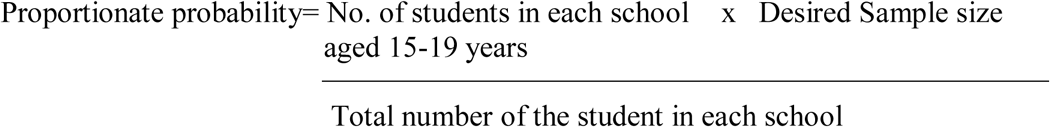

### Data Collection

The research instruments used were a pre-tested, semi-structured and self-administered questionnaire containing both open and closed ended questions. The questionnaire had different sections. The first section contained the socio-demographic characteristics of the respondents, the second section contained sexual behaviours and practice and the third section contained knowledge of sexual and reproductive health issues. Data were collected by trained research assistants and on different days of each school.

### Data Management

Data were analysed using descriptive statistics such as frequency, percentages, mean, and standard deviation with the use of Statistical Package for Social Sciences (SPSS version 21). Level of significance was considered at p < 5% for all inferential statistics.

### Ethical Considerations

Ethical approval was obtained from Joint University Of Ibadan/UCH Ethical Review Committee. Administrative approval was also obtained from the principals of each school selected for the study. Informed consent was obtained prior to the study. Participation in the study was voluntary and the questionnaire was administered by trained research assistants only. Privacy and confidentiality were ensured as the school teachers were not present during data collection, also, only serial number was used on the information collected rather than a name. The nature and purpose of the research and expectations from the students were duly explained to the students.

## RESULTS

This study included a total of 1227 female participants with age ranging between 15–19 years and mean age of 16.31±1.18 years. Table 1 shows that 33.7% of the respondent were from Osun central while 33.3% and 33% of the participant were from Osun west and Osun east respectively. The major religious affiliation among the respondent was Islam (73.1%), while (26.2%) was Christian and (0.6%) were traditional worshipper. Most of the respondents (99.4%) were single and only (0.6%) was married. Very Few (1.4%) of the respondents had a child and (98.6%) had not given birth at all. Most of the father’s (65.3%) of the respondents were self-employed, while (29.1%) and (5.6%) were employed and unemployed respectively. Majority of the mothers (79.4%) of the respondents were self-employed while about (16.5%) and (4.2%) were employed and unemployed respectively.

**Table 1:** Socio-demographics Characteristics.

| **Variables** | **Frequency (N)** | **Percentages (%)** |
| --- | --- | --- |
| **Senatorial District** |  |  |
| Osun central | 414 | 33.7 |
| Osun west | 408 | 33.3 |
| Osun east | 405 | 33.0 |
| **Religion** |  |  |
| Christian | 321 | 26.2 |
| Islam | 897 | 73.2 |
| Traditional | 7 | 0.6 |
| **Ethnicity** |  |  |
| Yoruba | 1218 | 99.4 |
| Igbo | 7 | 0.6 |
| **Do you have any children** |  |  |
| Yes |  |  |
| No | 17 | 1.4 |
|  | 1198 | 98.6 |
| **Father's occupation** |  |  |
| Unemployed | 68 | 5.6 |
| Employed | 353 | 29.1 |
| Self employed | 792 | 65.3 |
| **Mother's occupation** |  |  |
| Unemployed | 50 | 4.2 |
| Employed | 197 | 16.5 |
| Self-employed | 950 | 79.4 |
| **Education level of father** |  |  |
| No formal education | 82 | 6.7 |
| Primary education | 145 | 11.9 |
| Secondary education | 666 | 54.8 |
| Tertiary education | 322 | 26.5 |
| **Family type** |  |  |
| Monogamy | 627 | 52.2 |
| Polygamy | 575 | 47.8 |

Most of the respondent’s parent (79.5%) lives together while only about (20.5%) do not live together. More than half (52.2%) of the respondent were from a monogamous family while (47.8%) were from polygamous family.

Figure 1 shows the prevalence of sexual exposure of the respondents. 337(27.5%) were sexually exposed with a mean age of sexual debut 14.88±2.46 years.

**Figure 1:**
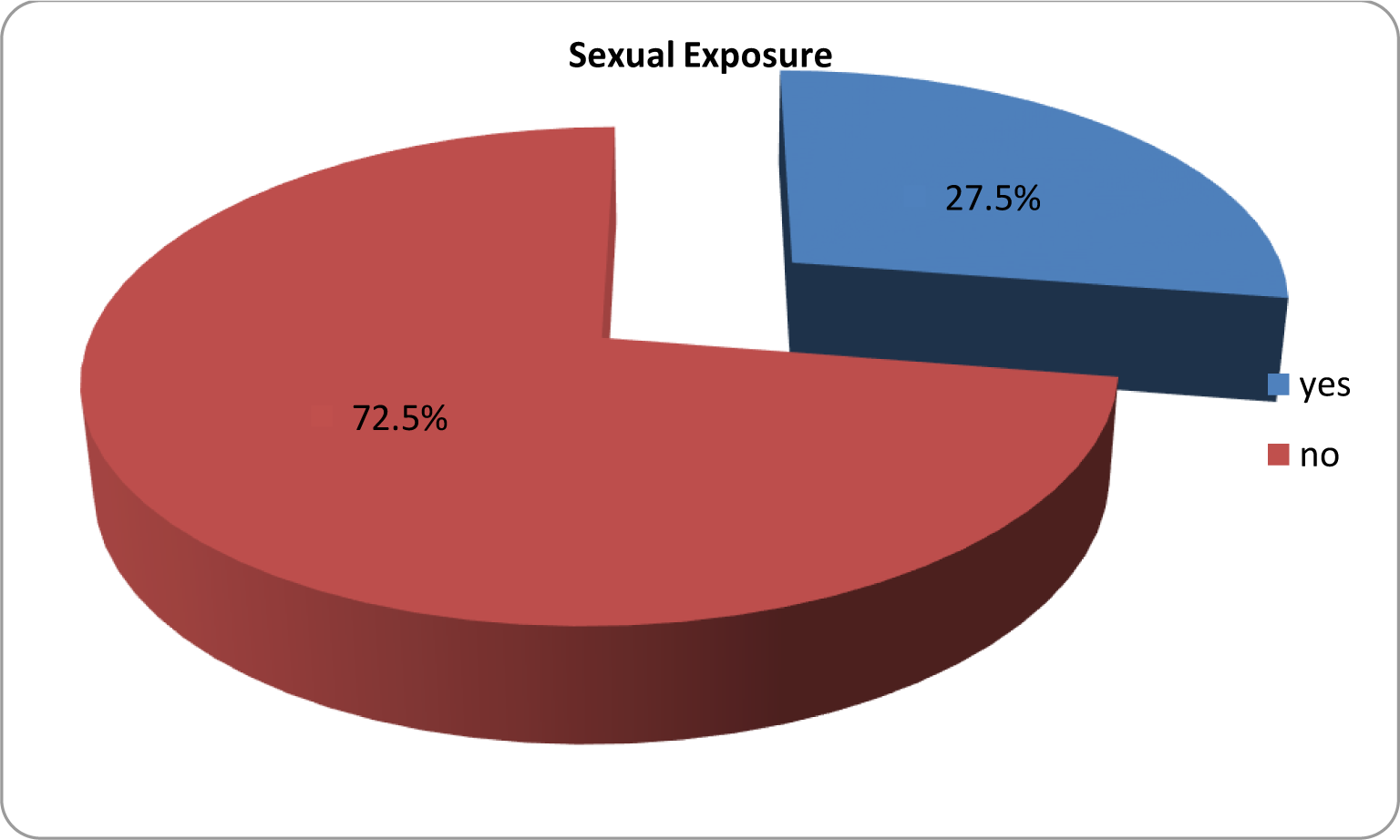
Sexual prevalence of the respondent.

### Sexual Behaviours and Practice

Table 2 shows the sexual behaviours and practice among the respondents. 337(27.5%) were sexually exposed with a mean age of sexual debut 14.88±2.46 years. Of the 337 that were sexually exposed 153(56.5%) initiated sex early between the ages of 10-15 years while 118(43.5%) initiated sex late between the ages of (16-19), sixteen (4.7%) had uses drugs or take alcohol before sexual intercourse while 315(95.3%) do not take alcohol before sexual intercourse.

**Table 2:** Sexual Behaviours and Practice.

| **Variables** | **Frequency** | **Percentage (%)** |
| --- | --- | --- |
| **Mean age at sexual initiation was** | **14.88±2.46** |  |
| **Have you ever had sexual intercourse** |  |  |
| Yes | 337 | 27.5 |
| No | 888 | 72.5 |
| **Sexual initiation** |  |  |
| Early initiation | 153 | 56.5 |
| Late initiation | 118 | 43.5 |
| **No of sexual partners in the last 12 month** |  |  |
| Single Partner | 250 | 81.2 |
| Multiple Partners | 58 | 18.8 |
| **Sexual intercourse with drugs or alcohol** |  |  |
| Yes | 16 | 4.7 |
| No | 315 | 95.3 |
| **Condom use with partner at last sexual intercourse** |  |  |
| Yes | 76 | 15.4 |
| No | 254 | 84.6 |
| **The last time you had sexual intercourse, what method** |  |  |
| **Did you use to prevent pregnancy** |  |  |
| No method was used to prevent pregnancy | 145 | 11.8 |
| Birth control pills | 30 | 2.4 |
| Condoms | 88 | 7.2 |
| Withdrawal method | 17 | 1.4 |
| Other method | 17 | 1.4 |
| No response | 63 | 5.1 |
| Not had sexual intercourse | 867 | 70.7 |
| **How many times have you been pregnant** |  |  |
| Once | 67 | 5.8 |
| twice | 2 | 0.2 |
| none | 1082 | 94.0 |
| **What happen to the pregnancy** |  |  |
| Currently pregnant | 12 | 1.0 |
| Abortion | 19 | 1.6 |
| Miscarriage | 15 | 1.3 |
| Live-birth(gave birth) | 23 | 2.0 |

Among those who those who had sex in the last 12 months preceding the study, 250(81.2%) had only one sexual partner, 37(12%) had two partners, 5(1.6%) had 5 partners, 7(2.3%) had four partners, 8(2.6%) and 1(0.3) had five partners and six partners respectively. Among respondents that were sexually exposed, 76(15.4%) practiced unprotected sex with their partner, while 254(84.6%) used protection during sexual intercourse. Also 145(11.8%) of the respondents who had been exposed to sex did not use any methods to prevent pregnancy, while 30(2.4%) used birth control pills to prevent pregnancy, 88(7.2%) used condoms, 17(1.4%) used withdrawal method and 17(1.4%) used other methods to prevent pregnancy.

Most of the respondents (94%) had not been pregnant, while 67 (5.8%) of the respondent had been pregnant once and 2(0.2%) twice. Of the 69 respondent who had pregnancy, 23(2%) gave birth, 19(1.6%) had abortion, 15(1.3%) had miscarriage and only 12(1%) were currently pregnant.

Figure 2 shows that among the reasons of exposure to sex, 107(9.6%) reported pressure from boyfriend/partner as reason for sexual initiation, 47(4.2%) reported peer pressure, 59(5.3%) indicated self-desire as a reason, 28(2.5%) reported curiosity, 17(1.5%) reported desire to get pregnant and 2(0.2%) said it was due to parent pressure.

**Figure 2:**
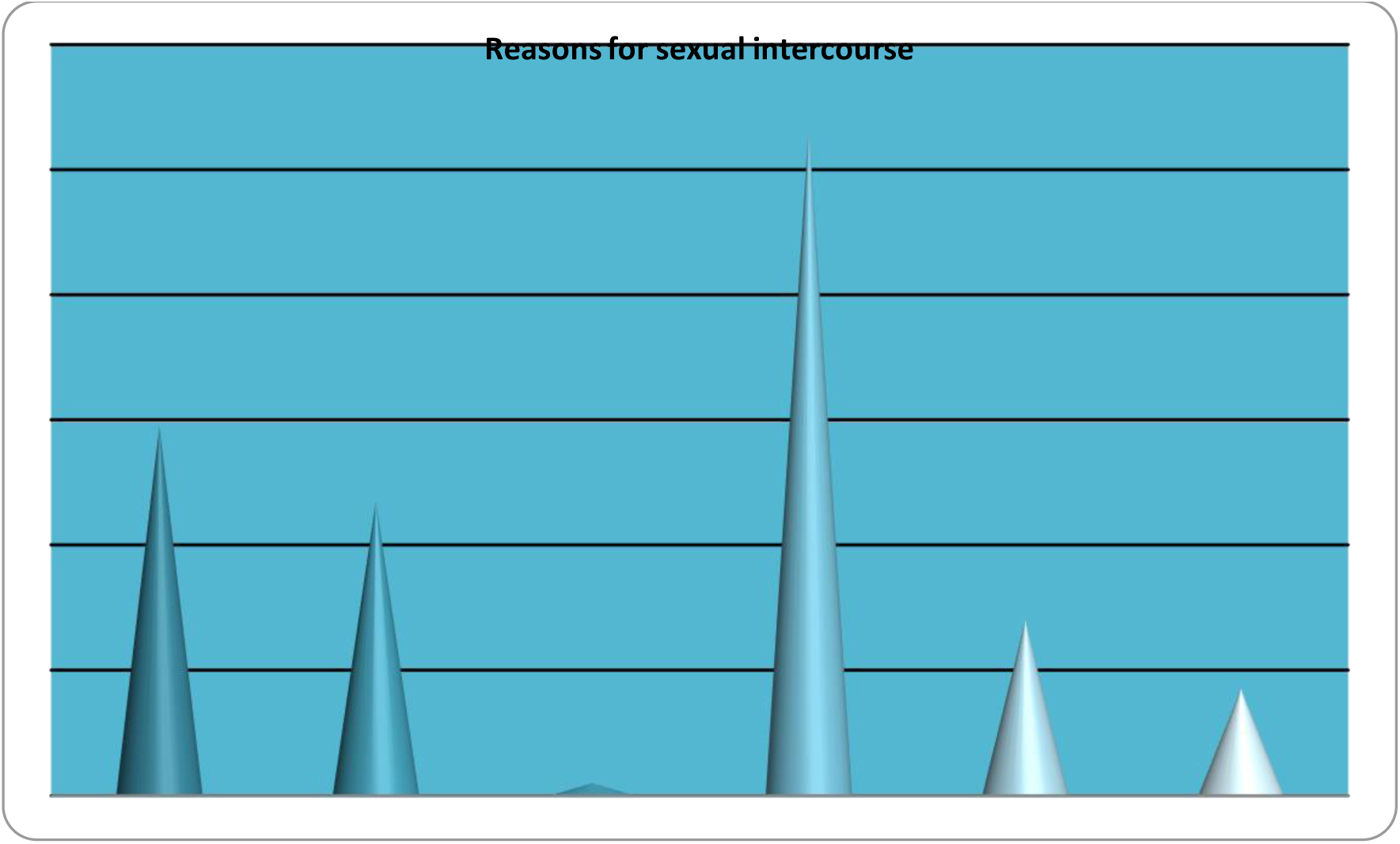
Reasons for sexual intercourse.

### Experience of sexual coercion

Table 3 shows that of the 337 respondents that were sexually exposed, 122(11%) were forced to have sex, 101(9.1%) played sex willingly, while 29(2.6%) felt threatened and 3(0.3%) were convinced with money. The proportion of the respondent who reported rape and abduction was (2.6%).

**Table 3:** Sexual Coercion.

| **Variables** | **Frequency (N)** | **Percentage (%)** |
| --- | --- | --- |
| **The first time you had sexual intercourse,**<br>**Did you agree willingly, or did it just happen**<br>**Or were you tricked, threatened or forced** |  |  |
| Played sex willingly (I wanted it) | 101 | 9.1 |
| Forced | 122 | 11 |
| Convinced with money or gift | 3 | 0.3 |
| Felt threatened | 29 | 2.6 |
| Not had sexual intercourse | 856 | 77.7 |
| **Have you any coercive sex practice like rape**<br>**and abduction** |  |  |
| Yes | 32 | 2.6 |
| No | 833 | 67.9 |
| No response | 362 | 29.5 |
| **What did you receive in exchange for sex** |  |  |
| Money | 41 | 3.5 |
| School fees | 24 | 2.0 |
| Shelter/rent | 2 | 0.2 |
| Gifts | 22 | 1.9 |
| Others | 35 | 3.0 |

Only 32(2.6%) of the respondent indicated they have been raped and abducted while 833(67.9) said they have not experience such things. Money was the highest 41 (3.5%) thing that have been received in exchange for sex, followed by school fees 24(2.0%). Also 22(1.9%) had received gift in exchange of sex while only 2(0.2%) received shelter in return for sex

## DISCUSSION

In this study socio-demographic characteristic of the participants showed that majority of the respondents were Muslims. All the respondents were aged between 15-19 years. Adolescent stage is a transition period to adulthood which is linked with series of issues that affect their body, behavior and social interactions (Duru *et.al*., 2010; Omobuwa *et.al*., 2012). The behaviours that are acquired during this transition period have great implications for their health and wellbeing and also they are likely to have proclivity for sexual experimentation and most of them initiate sexual activity during this period (Cherie and Berhane, 2012).

The findings of this study revealed that (27.5 %) of the respondents had had sexual intercourse before. This is in agreement with the findings by Asekun-Olarinmoye et al, in Osun State, Nigeria who reported that 27.6% of the in-school adolescents studied were sexually exposed (Asekun-Olarinmoye *et.al*.,2011) and Moharson-Bello *et al*. who reported 28.3% in a study carried out in Ibadan, Nigeria (Moharson-Bello *et.al*.2008).

In this study, the prevalence of sexually exposed adolescents is higher than the 13.0% reported among adolescents in Northeastern Nigeria (Ajuwon *et.al*., 2006) and South Africa (Peltzer, 2006). Higher prevalence rates were reported in studies by Slap *et al*. who reported 34% in Plateau State (Slap *et.al*.,2003), Owolabi et al. who reported 63% in Osun State (Owolabi *et.al*., 2005) and Olugbenga-Bello *et al*. who reported 31.5% also in Osun State, Nigeria (Olugbenga-Bello *et.al*., 2009). This variation in the proportion of adolescents who have been involved in sexual intercourse agrees with the findings in a review of studies on the sexual behavior of school students in sub-Saharan Africa with Nigeria inclusive, the prevalence of first sex was reported to vary from 3% to 93% (kaaya *et.al*.,2002). This variation may be due to the different values attached to the issue of adolescent sexuality by different people and people.

The mean age at sexual at first sexual intercourse was 14.88±2.46 years, this is similar to the findings by (Duru *et.al*., 2010 and Olugbenga-Bello *et.al*., 2009) who reported mean ages at sexual debut of 15.08±0.2, 15.2±1 years respectively in Nigeria. Other studies by (Masatu *et.al*., 2009; Chartsbin, 2008) conducted among in-school adolescents in Tanzania showed mean age at sexual debut to be 14.6 and 15.5 years respectively.

The major reasons for sexual exposure was pressure of partner/boyfriend and self-desire, this agrees with (Kora and Haile 2007) who noted that the most common reasons for youths experimenting sex before their 18th birth day include personal desire and peer pressure.

Of those that had sexual intercourse in the last 12 month preceding the study, about 19% of the respondents had had sexual intercourse with more one partner, and 10.6% had received money, school fees, shelter, others (such as different types of gifts) in exchange for sex. 4.7% out of the 27.5% that had had sexual exposure took drugs/alcohol before sex. This pattern of high risky behaviours has been well established by precious studies, though the findings vary from place to place.

A finding in Osun state by (Olugbenga-Bello *et.al*., 2009) found that 14.6% of the secondary school students surveyed had more than one sexual partner while another study found out in Osun state that of the sexually active students that were surveyed, 48.4% had multiple sexual partners (Asekun-Olarinmoye *et.al*.,2011). Also a study carried out among secondary schools student in Ilorin, Kwara state found out that 24.2% of the respondents had received gifts in exchange for sex, while 45% had more than one sexual partner. This high levels of risky sexual behavior have made adolescents particularly vulnerable to and at risk of STI and HIV infections, and hence, the reason why evidenced-based interventions should be directed at this age group.

From this study, out of those who had had sexual exposure, it is something of concern that just only (15.4%) used condom for the last sexual experience and this is similar with other findings that reported this poor preventive culture among adolescents. In a study conducted in Osogbo, Southwestern Nigeria, it was found that less than a third of the sexually active adolescents used condoms by (Olugbenga-Bello *et.al*., 2009) and while Nigeria Demographic and Health Survey reported that among young people that had sexual intercourse in the 12 months preceding the survey, 94.5% of male adolescents had high-risk intercourse and only 36.3% of them used condom (NDHS, 2008).

This low rate of condom use among respondents may be due to poor comprehensive knowledge about contraception, STIs and related issues among adolescents (Olugbenga-Bello *et.al*., 2010; Asekun-Olarinmoye *et.al*.,2011; Oyo-Ita *et.al*., 2005).

Teenage pregnancy is one of the most unfavorable and usually unplanned outcomes of adolescent sexual activity. In this study about 6% of the respondents had been pregnant, this is a little higher than the findings of (Olugbenga-Bello *et.al*., 2014) in Osun state that found that 4.3% had been pregnant, while the majority of those who had been pregnant gave birth and had had an abortion. Having an abortion has a lot of implications for reproductive health including septic abortions, damage to reproductive organs, blood borne infections, uterine perforations, and mortality. In addition, having a baby while in school may lead to school drop-out, and reduced opportunities to continue schooling, poor cohesion among families, rejection by the family and society, lower education, early marriage, and soaring fertility.

## CONCLUSION

Finding from this study is consistent with earlier studies conducted in many other Nigerian cities which showed that in-school adolescents to be sexually active. In fact, one can conclude from this study that at least one out of every four in-school adolescents in Osun state is sexually active and most engaged in unsafe sexual practices, therefore, this makes them vulnerable to various medical complications including STIs, HIV/AIDS and genital cancers. There is the need to step up campaigns to address this noticed lapse in behavior among the students in order to arrest the usual consequences of such risky sexual behavior. Interventions including peer education, training of teacher’s and development of youth friendly centers are recommended to meet the reproductive health needs of adolescents

## Data Availability

Data is archived in my data bank

